# Trends in the Association Between Meeting the Physical Activity Guidelines and Risk of Total and Cause-specific Mortality in the US Over 2 Decades

**DOI:** 10.1101/2023.09.08.23295287

**Authors:** David Martinez-Gomez, Fernando Rodriguez-Artalejo, Ding Ding, Ulf Ekelund, Veronica Cabanas-Sanchez

## Abstract

**Background:** It is unclear whether the association between meeting the physical activity (PA) guidelines and mortality varied in the last two decades among the US adult population.

**Methods:** We included seventeen annual representative samples of US adults in the National Health Interview Survey (1998-2014; n=482,756 individual participant data) and all-cause and cause-specific mortality ascertained through December 2019. Participants were grouped according to the 2018 US PA Guidelines: 150 or more min/week in aerobic PA and muscle-strengthening activities 2 or more times/week. To provide further context, we also examined the trends in mortality risk associated with other modifiable health factors.

**Results:** Meeting the PA guidelines was associated with lower 5-year all-cause mortality risk (HR=0.59, 95%CI, 0.55, 0.63) based on the pooled analyses, but there was a nonsignificant trend association (P for trend=0.305) between meeting PA guidelines and 5-year mortality across the seventeen annual surveys. Meeting the recommended PA guidelines was associated with 5-year cardiovascular (HR=0.59, 95%CI, 0.50, 0.69) and cancer mortality (HR=0.76, 95%CI, 0.64, 0.90) without indication of trend in the associations (P for trend=0.975 and 0.366, respectively). In pooled analyses, attain a high educational level, body mass index less than 30 kg/m^2^, being noncurrent smoker, nonheavy drinker, and living without history of hypertension and diabetes with 5-year mortality were 0.70 (95%CI, 0.67, 0.73), 1.19 (95%CI, 1.15, 1.23), 0.56 (95%CI, 0.54, 0.59), 0.85 (95%CI,0.79, 0.92), 0.91 (95%CI, 0.88-0.94) and 0.65 (95%CI, 0.88, 0.94), respectively, but only diabetes showed a significant trend analysis (P for trend<0.001) that attenuated the association.

**Conclusions:** Meeting PA guidelines lower mortality risk and this association has not changed in the US adult population over the last two decades. Encouraging adults to meet the PA guidelines may provide substantial health benefits, despite social, demographic and lifestyle changes, as well as the advances in medical technology and pharmacological treatments.

**Clinical Perspective:** *What is new?:*

- It is unclear whether the social, demographic, and lifestyle changes, well as the advances in medical technology and pharmacological treatments in the last two decades have influenced the relationship between the recommended physical activity guidelines and the risk of mortality.
- In this observational study that included 482 756 individual-participant data from 17 annual nationwide surveys, the association between meeting physical activity guidelines and all-cause and cause-specific mortality risk has not changed, that is, remained stable, in the US adult population.
- We provided context for our findings examining the trends in mortality risk associated with other modifiable health factors and only living without diabetes mellitus showed that the association was attenuated over the study time.

*What Are the Clinical Implications?:*

- These findings reinforce the public health message of the 2018 Physical Activity Guidelines by encouraging Americans to be physically active to provide substantial health benefits for the adult population.

## INTRODUCTION

Compelling evidence suggests that regular physical activity prevents premature mortality (1,2). To obtain this and others health benefits, the 2018 US physical activity guidelines (3) suggest that adults do at least 150 minutes a week of moderate-intensity, 75 minutes a week of vigorous-intensity, or an equivalent combination of moderate- and vigorous-intensity aerobic physical activity. Muscle-strengthening activities on 2 or more days a week are also recommended. Recent surveillance data suggest a positive trend in the prevalence of adults meeting the physical activity guidelines in the US (4,5). From 1998 to 2018, the prevalence of meeting the aerobic guidelines increased from 40% to 54% (4), meeting the muscle-strengthening guidelines increased from 18% to 28% (5), and meeting the combined aerobic and muscle-strengthening guidelines increased from 14% to 24% (5).

Besides changes in physical activity patterns, the US population has also undergone other changes in lifestyle risk factors. For example, the prevalence of adult cigarette smoking has steadily declined (6), whereas alcohol consumption, specially binge drinking behavior, has increased (7,8). The prevalence of adult obesity has increased (9) simultaneously with an improvement in diet quality (10). These changes in lifestyle risk factors coincide with demographic changes including increasingly diverse ethnic composition (11).

While the US population is characterized by rapid aging (12), mortality rates have declined by 17% and life expectancy increased by 2 years from 1998 to 2018 (13); the corresponding decreases in cardiovascular and cancer mortality were 40% and 26%, respectively. This seemingly paradox may partly be explained by major medical advances in the last decades in disease prevention, detection, and treatment of major chronic conditions (e.g., cardiovascular disease and cancer), and the management of modifiable health factors (e.g., quit tobacco, losing weight, high blood sugar and blood pressure control).

It is unclear how the changes of the last decades have influenced the relationship between the recommended physical activity guidelines for the US population and the risk of mortality. Thus, the aim of this study was to examine the trends (i.e., variations over time) in the association between meeting the physical activity guidelines and mortality in annual nationwide US populations from the last two decades. To provide a context for our findings, we also examined the trends in mortality risk associated with other health factors, such as smoking and obesity in these populations.

## METHODS

### Study design and participants

Data for this study are from the US National Health Interview Survey (NHIS). The NHIS is a survey annually conducted by the National Center for Health Statistics (NCHS), which is part of the US Centers for Disease Control and Prevention (CDC). The NHIS uses a multi-stage probability sampling designed to collect nationally representative estimates of the US civilian, noninstitutionalized population, and has monitored the health of the nation since 1957. Data are currently collected through in-person household computer-assisted personal interviewing, with telephone follow-up when the interview cannot be completed in person. Further details on this survey are available on the NHIS website (https://www.cdc.gov/nchs/nhis/). All participants provided informed consent before participation, and NHIS procedures were approved by the NCHS Research Ethics Review Board. No additional institutional approval was required to conduct secondary analyses using NHIS public-use data.

For this work we selected NHIS surveys from 1998 to 2014 (Figure 1). Since the NHIS protocols underwent major revisions in 1997 and 2019, we did not include surveys from before 1997 or after 2018. The 1997 cohort was excluded because the physical activity questionnaire was not administered in the same format throughout the year. The cohorts from 2015 to 2018 were also excluded because the last available date ascertained for vital status (December 31, 2019) did not allow a minimum of 5 years follow-up for these participants. In each cohort, we also excluded participants lacking data on physical activity variables, pregnant women at interview time, and those without information on vital status. Finally, a total of 482 756 participants from 17 annual surveys, aged 18 years or older, met the enrolment criteria and were included in this study.

**Figure 1.**
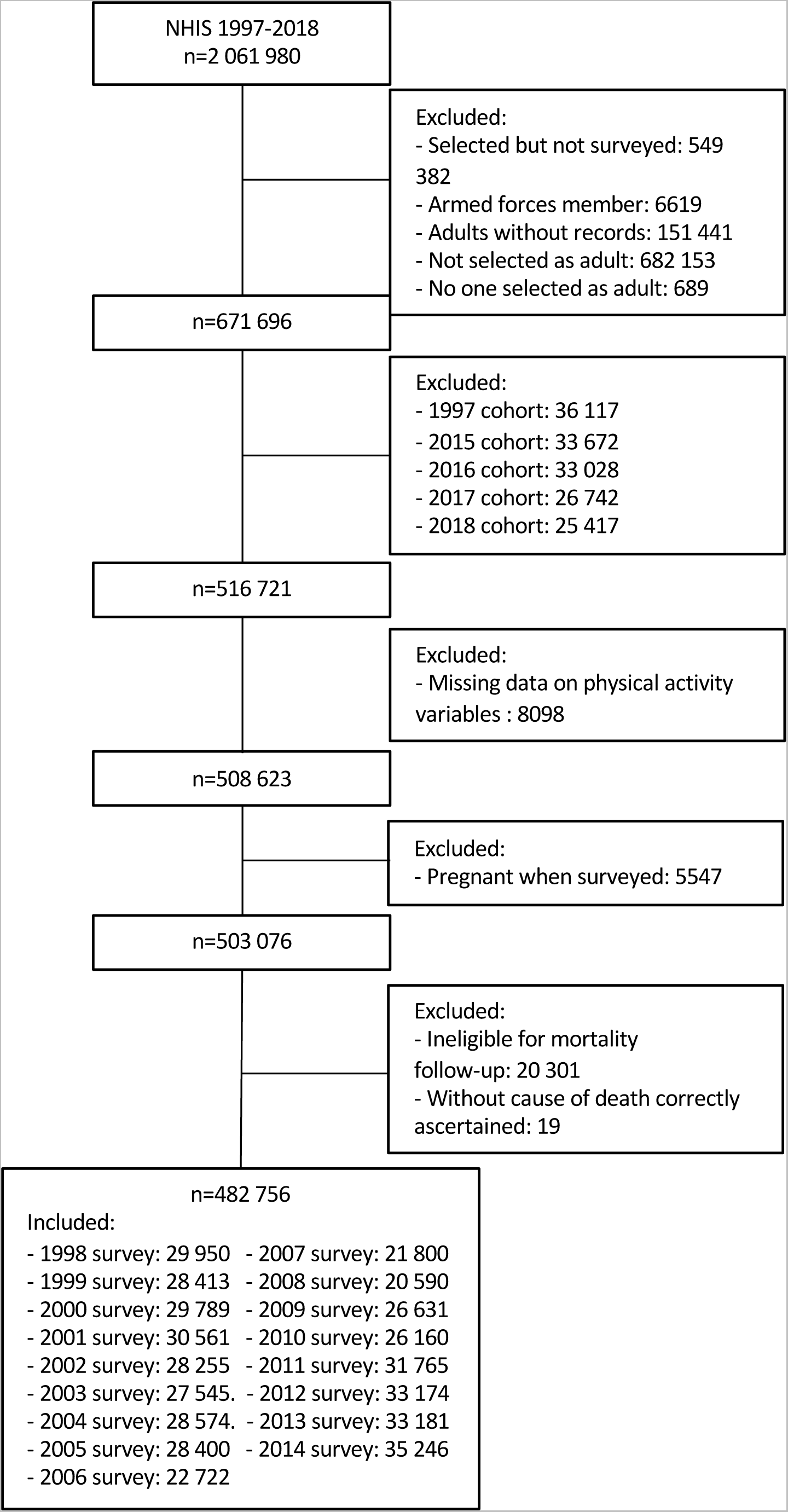
Participants flow diagram.

### Physical activity

Participants were asked how often and, if applicable, the duration of aerobic activities during leisure-time in i) vigorous (i.e., heavy sweating or large increases in breathing or heart rate) and ii) light-to-moderate (i.e., light sweating or slight to moderate increases in breathing or heart rate) levels (4,5). The total amount of aerobic physical activity was calculated by summing the minutes of vigorous-intensity (equivalent to around 6 metabolic equivalents) activity weighted by a factor of 2 and light-to-moderate-intensity activity (equivalent to around 3 metabolic equivalents). Muscle strengthening activity was assessed with the following question (5): “How often do you do physical activities specifically designed to strengthen your muscles such as lifting weights or doing calisthenics?”. The physical activity guidelines for adults recommend at least 150 minutes per week in aerobic physical activity and participating in muscle-strengthening activities 2 or more times per week, and hence, participants were categorized as meeting the physical activity guidelines if they reported meeting both recommendations (3).

### Mortality

Leading underlying cause of death was determined by NCHS based on probabilistic matches of survey participants’ NHIS records to National Death Index and classified using the 10th revision of the International Statistical Classification of Diseases, Injuries, and Causes of Death (ICD-10) (14). The accuracy of information on cause of death mortality in the National Death Index records has been validated in previous works (15,16). In the present study and to minimize bias owing to different follow-up durations, we calculated 5-year all-cause mortality for all participants. Deaths due to cardiovascular (I00-I09, I11, I13, I20-I51, I60-I69) and cancer (C00-C97) diseases, the leading causes of death in US, were also included as secondary study outcomes.

### Other study variables

Additional study variables were also reported during the interview. Sociodemographic factors included sex (male, female), age (years), race and ethnicity (non-Hispanic White, non-Hispanic Black, Hispanic, other), marital status (married or living with a partner, divorced, separated, or widowed, never married), and educational attainment (less than high school, high school grade or equivalent, and more than high school). Self-reported medical diagnoses of chronic and cardiometabolic conditions such as heart disease (yes, no), stroke (yes, no), respiratory disease (yes, no) including chronic bronchitis, asthma, or emphysema, hypertension (yes, no), and diabetes (yes, no) were also obtained in the survey. People taking blood pressure-lowering or antidiabetic medications, including insulin, were also included as having hypertension or diabetes, respectively. Never, former, and current smokers were identified, and alcohol consumers were classified as never, former, light-moderate, and heavy if they drank ≥14 drinks/week for men and ≥10 drinks/week for women during the past year. Self-reported weight and height was used to calculate the body mass index (kg/m^2^). Since self-reported body mass index values are subject to measurement error (e.g., tendency to over-report height and under-report weight), we initially adjusted these values with the Stommel & Schoenborn’s equation (17,18) derived from measured and self-reported weight and height data taken from the National Health and Nutrition Examination Survey 2001-2006, and after this adjustment, participants were classified as non-overweight/obesity (<25.0 kg/m^2^), overweight (25.0-29.9 kg/m^2^), and obesity (≥30.0 kg/m^2^). Missing data in each of these variables (due to participants not being sure, not knowing the answer, or refusing to respond), always lower than 5%, were handled using a missing indicator approach.

### Statistical analysis

Baseline characteristics of the study sample by NHIS cohort are presented as frequency (%). Cox regression was used to estimate hazard ratios (HR) and 95% confidence interval (CI) for the pooled association (i.e., all cohort as a unique cohort) between meeting physical activity guidelines (no vs. yes) and 5-year all-cause mortality across the seventeen cohorts from 1998 to 2014. To examine the trends in these associations, we initially calculated the HR and 95%CI for the association between meeting the physical activity guidelines and 5-year all-cause mortality separately for each cohort and thereafter summarized the time trend analysis with standardized B and P for trend values obtained from linear regressions, that used the HR and survey year as dependent and independent variables, respectively. We examined if the main results varied with cause-specific mortality due to cardiovascular and cancer diseases. For this cause-specific mortality analysis, we also excluded participants with prevalent cardiovascular disease for the cardiovascular mortality analysis and with prevalent cancer for the cancer mortality analysis. All Cox regression models were adjusted for sex, age, ethnicity and race, educational attainment, marital status, smoking, alcohol consumption, body mass index, heart disease, stroke, cancer, respiratory disease, hypertension, and diabetes. These main analyses were also stratified by nonmodifiable characteristics that have changed in the US society, including sex, age, ethnicity and race, and chronic conditions (healthy people without chronic conditions and people with one or more chronic conditions; cardiovascular disease, cancer, and respiratory disease were also specifically considered for stratification).

Additional analyses examined i) the dose-response association between four combined categories of meeting physical activity guidelines (i.e., insufficient aerobic and muscle strengthening activities, insufficient aerobic activity and recommended muscle strengthening activity, recommended aerobic activity and insufficient muscle strengthening activity, recommended both aerobic and muscle strengthening activities) and 5-year mortality, ii) the mutually adjusted associations of meeting the aerobic physical activity and muscle-strengthening recommendations with 5-year mortality, and iii) the dose-response associations of amounts of aerobic physical activity (i.e., 0 min/week, 1.0-149.9 min/week, 150-300 min/week, and >300 min/week) and muscle-strengthening activity (0 times/week, 0.1-1.9 times/week, 2.0-3.9 times/week, and ≥4 times/week) with 5-year mortality.

To place our trend analysis in context, we conducted a further analysis to investigate the association of modifiable health factors with 5-year mortality in pooled and time trend analysis. Among the health factors we selected: i) attain a high educational level (more than high school), ii) manage weight (body mass index <30 kg/m^2)^, iii) noncurrent smoker, iv) nonheavy drinker, v) manage blood pressure (never diagnosed with hypertension neither taking related medication), and vi) manage blood sugar (never diagnosed with diabetes neither taking related medication).

Finally, to account with a larger follow-up and consequently a greater number of deaths in each cohort year, we performed main analyses using 10-year mortality restricted to NHIS cohorts from 1998 to 2009 as a sensitivity analysis.

All analyses accounted for the complex survey design of NHIS by considering sample weights, and primary sampling units and stratum for variance estimation. All analyses were conducted using STATA version 14.2 (Stata Corp, College Station, TX, USA), with level of significance set at two-sided p<0.05.

## RESULTS

Baseline characteristics of the analytical sample included in this study are presented by survey year in Table 1. Overall, population in recent surveys are older, with a greater number of Hispanic people, more educated, never been married, never smokers, and with a higher prevalence of cancer, respiratory diseases, hypertension, and obesity. Adults in recent surveys seem to be more physically active since a greater percentage of people meet the physical activity guidelines for adults.

**Table 1.** Baseline characteristics of the analytical sample from the US National Health Interview Surveys 1998-2014.

|  | 1998 | 1999 | 2000 | 2001 | 2002 | 2003 | 2004 | 2005 | 2006 | 2007 | 2008 | 2009 | 2010 | 2011 | 2012 | 2013 | 2014 |
| --- | --- | --- | --- | --- | --- | --- | --- | --- | --- | --- | --- | --- | --- | --- | --- | --- | --- |
| *n* | 29950 | 28413 | 29789 | 30561 | 28255 | 27545 | 28574 | 28400 | 22722 | 21800 | 20590 | 26631 | 26160 | 31765 | 33174 | 33181 | 35246 |
| **Sex, %** |  |  |  |  |  |  |  |  |  |  |  |  |  |  |  |  |  |
| Female | 51.4 | 51.5 | 51.5 | 51.5 | 51.2 | 51.4 | 51.4 | 51.3 | 51.3 | 51.2 | 51.2 | 51.2 | 51.2 | 51.0 | 51.3 | 51.4 | 51.3 |
| **Age, %** |  |  |  |  |  |  |  |  |  |  |  |  |  |  |  |  |  |
| 18-39 years | 43.1 | 42.5 | 42.1 | 41.5 | 40.8 | 40.5 | 40.1 | 39.7 | 39.3 | 39.3 | 39.0 | 38.6 | 38.7 | 38.3 | 37.9 | 37.7 | 37.5 |
| 40-59 years | 34.9 | 35.6 | 36.2 | 36.9 | 37.2 | 37.3 | 37.2 | 37.6 | 38.0 | 37.1 | 37.4 | 37.3 | 36.5 | 36.2 | 36.2 | 35.9 | 35.5 |
| 60 or more years | 22.0 | 21.9 | 21.7 | 21.6 | 22.0 | 22.2 | 22.6 | 22.7 | 22.6 | 23.6 | 23.5 | 24.1 | 24.7 | 25.4 | 25.9 | 26.4 | 27.0 |
| **Race or ethnicity, %** |  |  |  |  |  |  |  |  |  |  |  |  |  |  |  |  |  |
| Non-Hispanic white | 75.5 | 75.1 | 74.4 | 73.9 | 73.7 | 72.8 | 72.7 | 72.2 | 70.3 | 69.7 | 69.3 | 69.1 | 68.7 | 68.4 | 67.2 | 66.8 | 66.4 |
| Non-Hispanic black | 11.0 | 11.0 | 11.2 | 11.2 | 11.2 | 11.0 | 11.1 | 11.3 | 11.5 | 11.6 | 11.6 | 11.7 | 11.8 | 11.7 | 11.7 | 11.8 | 12.0 |
| Hispanic | 9.8 | 10.1 | 10.4 | 10.6 | 10.8 | 12.1 | 12.1 | 12.2 | 13.0 | 13.2 | 13.5 | 13.7 | 14.0 | 14.1 | 14.9 | 15.1 | 15.3 |
| Other | 3.7 | 3.7 | 4.0 | 4.3 | 4.3 | 3.9 | 4.1 | 4.2 | 5.1 | 5.4 | 5.3 | 5.2 | 5.4 | 5.7 | 6.0 | 6.1 | 6.2 |
| **Education, %** |  |  |  |  |  |  |  |  |  |  |  |  |  |  |  |  |  |
| < High school | 48.7 | 47.9 | 48.4 | 46.9 | 46.4 | 46.2 | 45.6 | 45.8 | 45.8 | 44.3 | 43.0 | 42.6 | 41.3 | 40.9 | 40.4 | 39.8 | 39.2 |
| High school or FED | 28.2 | 28.5 | 28.2 | 29.0 | 28.8 | 29.2 | 29.1 | 28.3 | 28.7 | 28.5 | 29.9 | 30.4 | 30.3 | 31.2 | 31.1 | 30.8 | 30.7 |
| > High school | 22.6 | 22.9 | 22.9 | 23.4 | 24.2 | 23.9 | 24.6 | 25.4 | 24.7 | 26.4 | 26.5 | 26.6 | 28.0 | 27.5 | 28.2 | 29.1 | 29.6 |
| **Marital status, %** |  |  |  |  |  |  |  |  |  |  |  |  |  |  |  |  |  |
| Married or living with a partner | 59.8 | 58.4 | 58.1 | 57.9 | 57.8 | 57.6 | 57.2 | 57.0 | 56.6 | 55.8 | 55.0 | 54.5 | 54.2 | 53.3 | 53.0 | 52.8 | 52.9 |
| Divorced/separated/widowed | 18.9 | 19.4 | 19.4 | 19.6 | 19.2 | 19.3 | 19.4 | 19.5 | 19.4 | 19.5 | 19.7 | 19.8 | 19.9 | 20.3 | 19.8 | 20.2 | 19.9 |
| Never married | 21.0 | 21.9 | 22.2 | 22.4 | 22.8 | 22.9 | 22.9 | 23.3 | 23.7 | 24.4 | 25.1 | 25.5 | 25.8 | 26.3 | 27.1 | 26.8 | 27.0 |
| **Chronic conditions, %** |  |  |  |  |  |  |  |  |  |  |  |  |  |  |  |  |  |
| Heart disease | 6.0 | 5.5 | 5.8 | 6.4 | 6.3 | 6.1 | 6.6 | 6.7 | 6.5 | 6.3 | 6.5 | 6.6 | 6.7 | 6.7 | 6.6 | 6.6 | 6.0 |
| Stroke | 2.3 | 2.2 | 2.3 | 2.4 | 2.4 | 2.5 | 2.6 | 2.5 | 2.7 | 2.5 | 2.9 | 2.7 | 2.8 | 2.7 | 2.8 | 2.8 | 2.7 |
| Cancer | 6.3 | 6.7 | 6.6 | 7.0 | 7.2 | 6.8 | 7.2 | 7.5 | 7.3 | 7.5 | 8.0 | 8.4 | 8.5 | 8.3 | 8.5 | 8.6 | 8.6 |
| Respiratory disease | 12.9 | 12.1 | 13.1 | 15.1 | 14.2 | 13.1 | 13.3 | 14.0 | 14.4 | 13.9 | 15.7 | 16.3 | 15.7 | 15.7 | 15.4 | 14.8 | 15.4 |
| **Cardiometabolic risk factors, %** |  |  |  |  |  |  |  |  |  |  |  |  |  |  |  |  |  |
| Hypertension | 23.3 | 23.1 | 23.0 | 24.1 | 24.8 | 25.7 | 26.0 | 26.4 | 27.5 | 27.8 | 29.6 | 29.3 | 30.5 | 30.1 | 30.1 | 29.9 | 31.4 |
| Diabetes | 5.4 | 5.5 | 6.0 | 6.5 | 6.8 | 6.8 | 7.2 | 7.7 | 8.4 | 8.4 | 8.8 | 9.7 | 9.8 | 9.6 | 9.7 | 10.1 | 9.9 |
| **Body mass index, %** |  |  |  |  |  |  |  |  |  |  |  |  |  |  |  |  |  |
| <24.9 kg/m<sup>2</sup> | 34.4 | 33.5 | 33.1 | 31.7 | 30.6 | 30.7 | 30.8 | 30.2 | 29.7 | 29.0 | 28.7 | 27.4 | 28.2 | 28.0 | 27.1 | 28.2 | 26.9 |
| 25.0-29.9 kg/m<sup>2</sup> | 36.1 | 35.6 | 35.3 | 35.6 | 35.7 | 35.6 | 35.2 | 34.8 | 33.8 | 34.3 | 34.3 | 34.5 | 33.9 | 33.9 | 34.2 | 32.9 | 33.8 |
| ≥30.0 kg/m<sup>2</sup> | 27.2 | 28.3 | 28.7 | 29.5 | 30.2 | 30.5 | 31.1 | 32.1 | 32.9 | 32.9 | 34.1 | 35.2 | 35.3 | 35.5 | 35.6 | 36.2 | 36.4 |
| **Smoking, %** |  |  |  |  |  |  |  |  |  |  |  |  |  |  |  |  |  |
| Never | 52.2 | 52.9 | 53.9 | 54.4 | 53.9 | 55.4 | 56.7 | 56.7 | 57.4 | 57.8 | 57.5 | 57.0 | 58.7 | 58.8 | 59.5 | 59.8 | 61.2 |
| Former | 23.1 | 23.1 | 22.3 | 22.4 | 23.0 | 22.1 | 21.8 | 21.8 | 21.3 | 21.8 | 21.6 | 22.1 | 21.7 | 21.9 | 22.1 | 22.1 | 22.0 |
| Current | 24.4 | 23.8 | 23.5 | 23.0 | 22.8 | 22.2 | 21.2 | 21.3 | 21.0 | 20.0 | 20.7 | 20.8 | 19.6 | 19.1 | 18.2 | 17.9 | 16.7 |
| **Alcohol consumption, %** |  |  |  |  |  |  |  |  |  |  |  |  |  |  |  |  |  |
| Abstainer | 21.1 | 21.9 | 23.5 | 21.9 | 21.3 | 23.8 | 23.6 | 23.2 | 24.4 | 22.9 | 20.6 | 19.4 | 20.4 | 19.6 | 20.6 | 20.7 | 20.9 |
| Former drinker | 15.8 | 14.7 | 14.2 | 14.7 | 15.0 | 14.1 | 14.3 | 14.2 | 14.0 | 14.7 | 14.4 | 14.9 | 14.3 | 14.6 | 14.3 | 14.0 | 13.5 |
| Light-moderate drinker | 58.4 | 58.6 | 57.9 | 58.2 | 58.5 | 57.0 | 56.6 | 57.1 | 55.7 | 56.4 | 59.3 | 60.3 | 60.4 | 61.1 | 60.3 | 60.1 | 60.6 |
| Heavy drinker* | 3.9 | 4.1 | 3.7 | 4.1 | 4.2 | 4.1 | 3.7 | 4.0 | 4.3 | 4.2 | 4.6 | 4.4 | 4.1 | 3.9 | 3.9 | 4.3 | 4.2 |
| **Meeting physical activity guidelines, %** |  |  |  |  |  |  |  |  |  |  |  |  |  |  |  |  |  |
| Aerobic guidelines | 40.7 | 41.3 | 42.5 | 43.3 | 43.2 | 43.9 | 42.0 | 41.1 | 41.2 | 41.4 | 43.5 | 46.7 | 46.7 | 48.2 | 49.5 | 49.2 | 49.3 |
| Muscle-strengthening guidelines | 18.2 | 18.4 | 18.4 | 20.5 | 20.7 | 20.9 | 19.9 | 20.3 | 19.5 | 19.9 | 21.9 | 22.4 | 23.9 | 23.9 | 23.7 | 23.9 | 24.2 |
| Both guidelines | 14.7 | 15.2 | 15.1 | 16.7 | 16.8 | 17.3 | 16.2 | 16.5 | 16.0 | 16.4 | 18.1 | 18.8 | 20.2 | 20.4 | 20.2 | 20.4 | 20.9 |

During the 5 years follow-up, a total of 27159 participants died, ranging from 1766 deaths in 1998 (5.9%) to 2213 in 2014 (6.3%). Meeting the physical activity guidelines was associated with lower 5-year mortality risk (HR=0.59, 95%CI, 0.55, 0.63) based on the pooled analyses (Figure 2). We consistently observed an inverse association in all years, but there was a nonsignificant trend association (B=0.26, 95%CI, −0.24, 0.66, P for trend=0.305) between meeting physical activity guidelines and 5-year mortality across the seventeen annual surveys. We rerun the main analyses with the secondary study outcomes of 5-year cardiovascular mortality (8805 cases) and cancer mortality (6742 cases) (Figure 3). Meeting the recommended physical activity guidelines was associated with 5-year cardiovascular (pooled HR=0.59, 95%CI, 0.50, 0.69) and cancer mortality (pooled HR=0.76, 95%CI, 0.64, 0.90). There was no indication of trend in the associations for these cause-specific mortality analyses (P for trend= 0.975 and 0.366, respectively).

**Figure 2.**
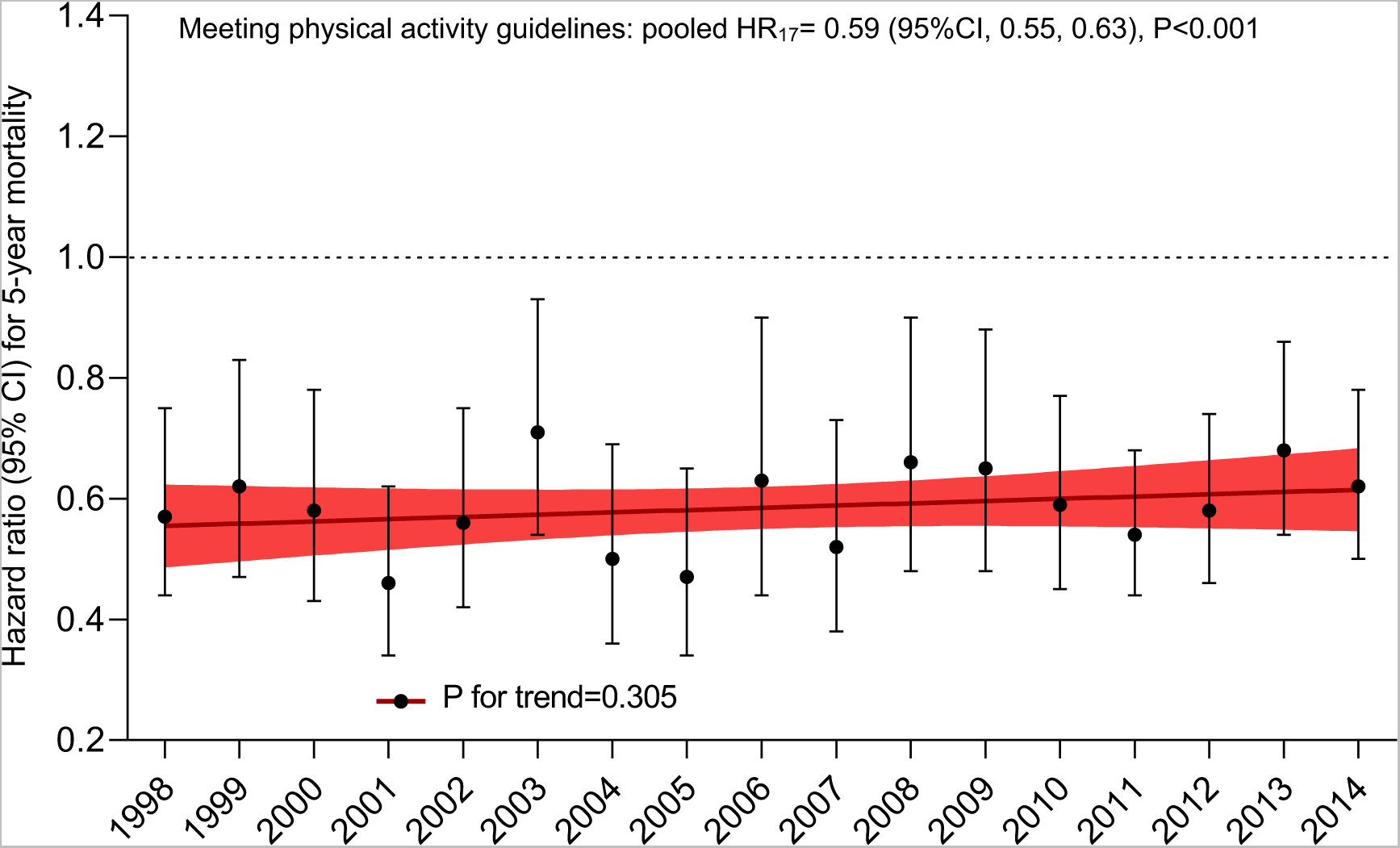
Associations between meeting physical activity guidelines and 5-year mortality in the National Health Interview Surveys 1998-2014. Pooled analysis included 17 annual surveys (1998-2014) as a unique cohort. Trend analysis was summarized with linear regression estimates (standardized B) that used the hazard ratio and survey year as dependent and independent variables, respectively. Meeting physical activity guidelines include both recommended aerobic and muscle strengthening activities. The color line indicates lineal regression and its 95%CI. Hazard ratios were adjusted for sex, age, race, education, marital status, chronic conditions, cardiometabolic risk factors, body mass index, smoking, and alcohol consumption.

**Figure 3.**
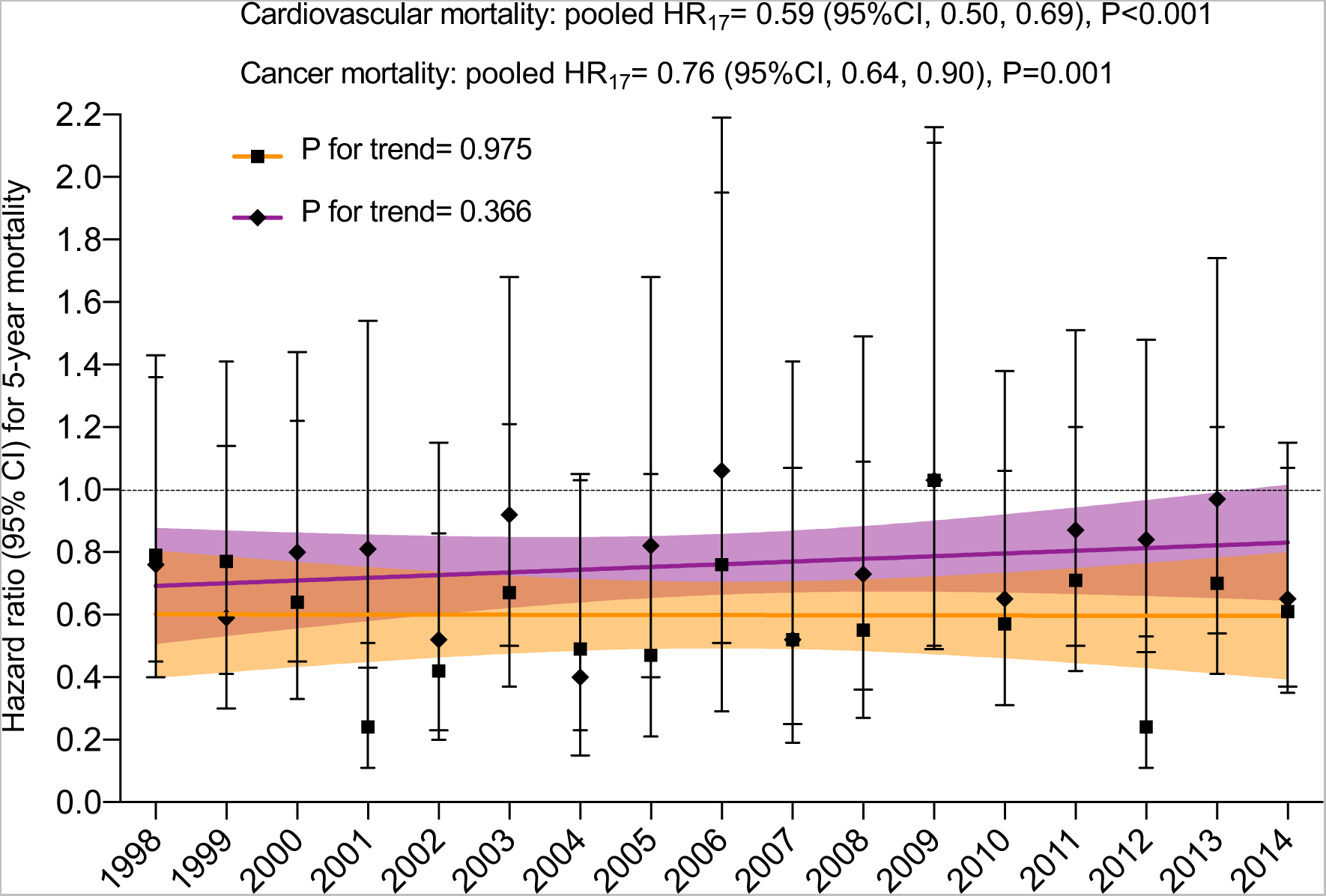
Associations between meeting physical activity guidelines and 5-year cardiovascular (orange) and cancer (purple) mortality in the National Health Interview Surveys 1998-2014. Pooled analysis included 17 annual surveys (1998-2014) as a unique cohort. Trend analysis was summarized with linear regression estimates (standardized B) that used the hazard ratio and cohort year as dependent and independent variables, respectively. Meeting physical activity guidelines include both recommended aerobic and muscle strengthening activities. The color lines indicate lineal regressions and its 95%CI. A total of 42527 participants with prevalent cardiovascular disease were excluded for the cardiovascular mortality analysis, and 38556 participants with prevalent cancer for the cancer mortality analysis. Hazard ratios were adjusted for sex, age, race, education, marital status, other chronic conditions, cardiometabolic risk factors, body mass index, smoking, and alcohol consumption.

Dose-response associations between the four combined groups of physical activity guidelines and 5-year mortality found similar results in pooled and trends analyses (Figure S1). Without exceptions, the pooled association between meeting physical activity guidelines with 5-year mortality were consistent when stratified by sex, age, ethnicity and race, and chronic conditions, without any trend associations (all P for trend >0.1) (Table 2).

**Table 2.** Associations between meeting physical activity guidelines and 5-year mortality stratified by nonmodifiable characteristics.

|  |  | Pooled analysis* |  | Trend analysis** |  |
| --- | --- | --- | --- | --- | --- |
|  | n/deaths | Hazard ratio (95%CI) | P | B (95%CI) | P for trend |
| **Sex** |  |  |  |  |  |
| Men | 214,526/13,122 | 0.61 (0.56, 0.67) | <0.001 | 0.30 (-0.20, 0.68) | 0.227 |
| Women | 254,193/14,037 | 0.54 (0.49, 0.60) | <0.001 | 0.02 (-0.45, 0.50) | 0.910 |
| **Age** |  |  |  |  |  |
| 18-59 years | 351,166/5506 | 0.67 (0.59, 0.76) | <0.001 | 0.12 (-0.37, 0.57) | 0.625 |
| ≥ 60 years | 131,590/21653 | 0.55 (0.51, 0.60) | <0.001 | 0.36 (-0.14, 0.71) | 0.151 |
| **Race or ethnicity** |  |  |  |  |  |
| Non-Hispanic white | 305,201/19,886 | 0.57 (0.53, 0.62) | <0.001 | 0.22 (-0.28, 0.63) | 0.222 |
| Non-Hispanic black | 69,981/4178 | 0.58 (0.48, 0.71) | <0.001 | -0.39 (-0.73, 0.10) | 0.118 |
| Hispanic | 82,308/2298 | 0.74 (0.57, 0.97) | 0.030 | -0.26 (-0.66, 0.24) | 0.306 |
| **Chronic conditions** |  |  |  |  |  |
| 0 chronic conditions | 355,104/11,284 | 0.70 (0.63, 0.77) | <0.001 | 0.04 (-0.44, 0.51) | 0.853 |
| ≥1 chronic conditions | 127,536/15,862 | 0.50 (0.45, 0.55) | <0.001 | 0.25 (-0.26, 0.65) | 0.332 |
| Cardiovascular disease | 42,523/9134 | 0.54 (0.47, 0.62) | <0.001 | 0.22 (-0.28, 0.63) | 0.380 |
| Cancer | 38556/6694 | 0.44 (0.39, 0.50) | <0.001 | 0.02 (-0.46, 0.50) | 0.917 |
| Respiratory disease | 70,911/5943 | 0.55 (0.47, 0.65) | <0.001 | 0.16 (-0.34, 0.59) | 0.527 |

Meeting physical activity guidelines include recommended both aerobic physical activity and muscle-strengthening activities. \* Pooled analysis with 17 annual surveys (1998-2014) as a unique cohort. \*\* Linear regression estimates (standardized B) that used the hazard ratio and survey year as dependent and independent variables, respectively. Hazard ratios were adjusted for sex, age, race, education, marital status, chronic conditions, cardiometabolic risk factors, body mass index, smoking, and alcohol consumption.

Meeting aerobic physical activity (HR=0.58, 95%CI, 0.56, 0.51) and muscle-strengthening (HR=0.86, 95%CI, 0.81, 0.90) guidelines were independently associated with 5-year mortality risk in pooled analyses, without any evidence for trends in the associations (P for trend= 0.448 and 0.360, respectively) (Figure 4). In mutually adjusted analyses (Figure S2), compared with the referent (i.e., no aerobic physical activity), risk reductions were 31%, 44% and 50% for 5-year mortality by increasing amounts of aerobic physical activity; the corresponding risk reductions for increasing categories of muscle-strengthening activities were 17%, 17% and 3%, independently of aerobic physical activity. The dose-response associations of amounts of aerobic and muscle-strengthening activities and mortality risk were stable during the study time (all P for trend >0.1).

**Figure 4.**
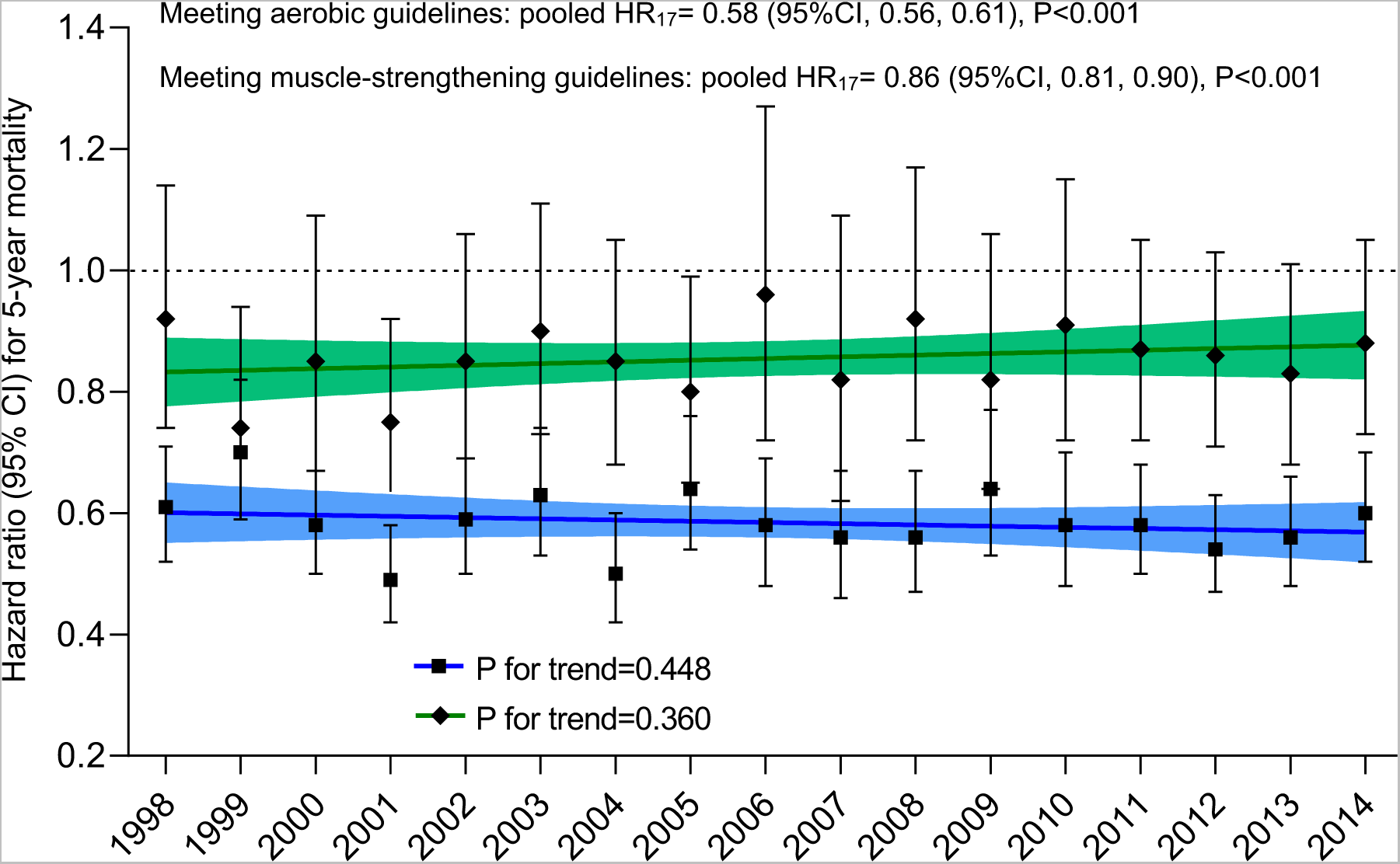
Associations between meeting aerobic (blue) or muscle-strengthening guidelines (green) and 5-year mortality in the National Health Interview Surveys 1998-2014. Pooled analysis included 17 annual surveys (1998-2014) as a unique cohort. Trend analysis was summarized with linear regression estimates (standardized B) that used the hazard ratio and cohort year as dependent and independent variables, respectively. The color lines indicate lineal regressions and its 95%CI. Hazard ratios were adjusted for sex, age, race, education, marital status, chronic conditions, cardiometabolic risk factors, body mass index, smoking, alcohol consumption, and meeting the other guideline.

Restricting our main results to cohorts with at least 10 years of follow-up (surveys from 1998 to 2009, n=323 230), and including a total of 38816 deaths, did not materially changed the results (HR=0.68, 95%CI, 0.65, 0.72 for the pooled analysis, and B=-0.11, 95%CI, −0.64, 0.48, P for trend=0.711, for the trend analysis) (Figure S3).

Finally, in pooled analyses (Figure 5), attain a high educational level, body mass index less than 30 kg/m^2^, being noncurrent smoker, nonheavy drinker, and living without history of hypertension and diabetes with 5-year mortality were 0.70 (95%CI, 0.67, 0.73), 1.19 (95%CI, 1.15, 1.23), 0.56 (95%CI, 0.54, 0.59), 0.85 (95%CI, 0.79, 0.92), 0.91 (95%CI, 0.88-0.94) and 0.65 (95%CI, 0.88, 0.94), respectively. Trend analyses of these health factors showed nonsignificant associations (all P for trend>0.4), with the exception of living without diabetes that showed a tendency (P for trend <0.001) that the association between living without diabetes and mortality were attenuated over the study time (B=0.77, 95%CI, 0.46, 0.91).

**Figure 5.**
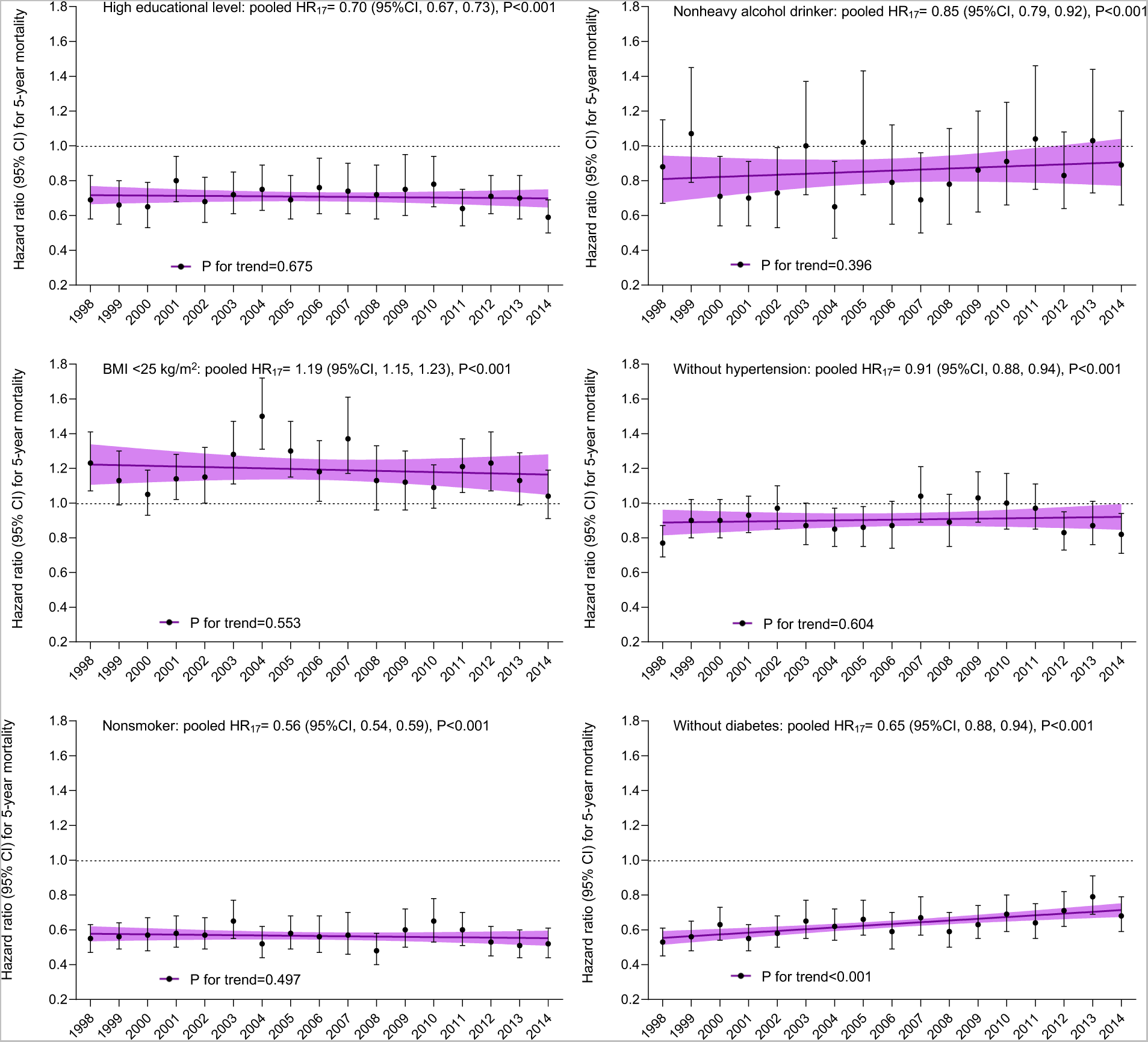
Associations between health factors and health behaviors and 5-year mortality in the National Health Interview Surveys 1998-2014. Pooled analysis included 17 annual surveys (1998-2014) as a unique cohort. Trend analysis was summarized with linear regression estimates (standardized B) that used the hazard ratio and cohort year as dependent and independent variables, respectively. High educational level: more than high school. Heavy drinker: ≥ 14 drinks/week for men and ≥ 10 drinks/week for women during the past year. The color line indicates lineal regression and its 95%CI. Hazard ratios were adjusted for sex, age, race, marital status, chronic conditions, meeting physical activity guidelines, and the other health factors.

## DISCUSSION

In the present study, we found that meeting the physical activity guidelines was associated with lower mortality risk in each of the seventeen annual nationwide cohorts of US adults from 1998 to 2014. The magnitude of the association remained stable over the study time, suggesting that the role of physical activity has always been important and remains important. These findings imply that all Americans should be physically active despite social, demographic and lifestyle changes in the US society in the last decades, as well as the advances in medical technology and pharmacological treatments.

A previous study by O’Donovan et al. (19), with a different aim that the one examined in the present study, showed the associations between meeting aerobic physical activity guidelines and all-cause mortality, stratified by survey and survey year as additional analyses. That study included eleven population-based British cohorts, eight from the Health Survey for England (1994, 1997, 1998, 1999, 2003, 2004, 2006, 2008) and three from the Scottish Health Survey (1995, 1998, 2003), including a total of 63 591 participants, aged 40 years or older. In agreement with the present study, in all cohorts, meeting aerobic physical activity guidelines lower all-cause mortality. Although interesting and allow us to provide certain data to compare our results, these results from the O’Donovan and colleagues’ work must be interpreted with caution because i) only age- and sex-adjusted HRs were included and hence, secular changes in sociodemographic, lifestyle, and health conditions in the British population could be not taken into account since only age- and sex-adjusted estimated were presented, ii) each cohort had different follow-up durations (mean=8.8, SD=4.4 years), which hamper direct comparisons across cohorts, and iii) physical activity data required certain harmonization due to slight differences by survey and changes over survey years.

Farrell et al. (20) examined whether the association between cardiorespiratory fitness, whose primary driver is physical activity levels, and mortality in an early cohort of men (1971 to 1991, n=24,475) persisted when evaluated in a more contemporary cohort of men (1992 to 2013, n=23,387) with identical follow-up periods. They found a significant inverse trend (P<0.001) between fitness levels and all-cause mortality in both cohorts (per 1-MET increment, HR=0.86, 95%CI, 0.82, 0.90 in cohort 1 and HR=0.87, 95%CI, 0.83, 0.91 in cohort 2). This study demonstrated that fitness also remains strongly associated with mortality in the current era of enhanced medical care.

In our study, mortality benefits of meeting physical activity guidelines were stable when main analyses were stratified by sex, but also by age, and race and ethnicity; two sociodemographic conditions that have changed over time in the US nation. Also, we hypothesized that among people with chronic conditions who requires medical care (21), the beneficial effect on survival associated with physical activity will decrease over time because of improvements in health care and disease management which prolong life; however, we observed stability in all these associations.

As expected, meeting the aerobic physical guidelines reduced the mortality at greater extent than meeting muscle-strengthening activity guidelines, but both types of physical activity showed analogous benefits over time. The mortality risk reductions of different amounts of aerobic and muscle-strengthening physical activities were also stable over time; when we could think that lower amounts of physical activity would lose benefits due to medical advances and higher amounts would be needed to maintain them. Finally, to put our results in context, we examined the trends in the associations of other health metrics with mortality risk, including six modifiable health factors. This further analysis also found stability in the mortality benefits associated to healthier factors over time, with the exception of living with diabetes, that showed an attenuation in the association (P for trend <0.001). Mortality benefits of living without diabetes as compared to people with diabetes among US people seem to be reducing every year. Improvements in early detection and development and accessibility to new medications could be playing an important role in this result that, albeit positive, warrants further research.

The strengths of the present study include the use of large, annual, and nationally representative cohorts of the US population. In addition, and importantly, all study variables were measured with the same methods across the surveys. However, some study limitations should be considered. First, information on physical activity was self-reported and, hence, prone to information bias from recall and social desirability (e.g., tendency to overreporting). To reduce these limitations, more objective methods to assess physical activity (e.g., accelerometers) may be considered in future studies. Second, we only had a single physical activity assessment at baseline; repeated measures of the exposure variable, at least, could have minimized measurement error and within-individual variability during the follow-up (22). These potential reporting and measurement biases would likely have led to an underestimation of the observed effect sizes of physical activity on mortality in our work (22). Third, although the analyses were adjusted for sociodemographic, lifestyle, and health-related conditions, residual confounding is still likely due to unmeasured (e.g., diet and subclinical diseases) and poorly measure confounders (e.g., self-reported weight and height, and alcohol). Fifth, this was an observational study, which limits causal inference. Finally, the other health factors analyzed in our study were also based on self-report. Thus, there may also be prone to the form of biases as physical activity data. Further research should consider including more accurate measurements (e.g., clinical blood pressure monitoring, fasting glucose or glycosylated hemoglobin in blood samples, measures of body weight with a balance and stature with a stadiometer, accelerometers) and repeated measures for these health factors to confirm our results.

In conclusion, the association between meeting physical activity guidelines and mortality risk has not changed in the US adult population over time. Encouraging adults to meet the physical activity guidelines may provide substantial health benefits for the US population. The findings can be used to reinforce the public health message of the 2018 and future US physical activity guidelines.

## Author Contributions

DMG and VCS had full access to all of the data in the study and take responsibility for the integrity of the data and the accuracy of the data analysis.

Concept and design: DMG, VCS. Acquisition, analysis, or interpretation of data: All the authors. Drafting of the manuscript: DMG, UE, DD. Critical revision of the manuscript for important intellectual content: All authors. Statistical analysis: DMG, VCS. Obtained funding: FRA, DMG. Administrative, technical, or material support: FRA, VCS. Supervision: FRA, UE, DD.

## Disclaimer

The findings and conclusions in this article are those of the authors and do not necessarily represent the official position of the NCHS/CDC and funders/sponsors.

## Data Availability

Data is available in https://www.cdc.gov/nchs/nhis/index.htm

---

**Figure S1.**
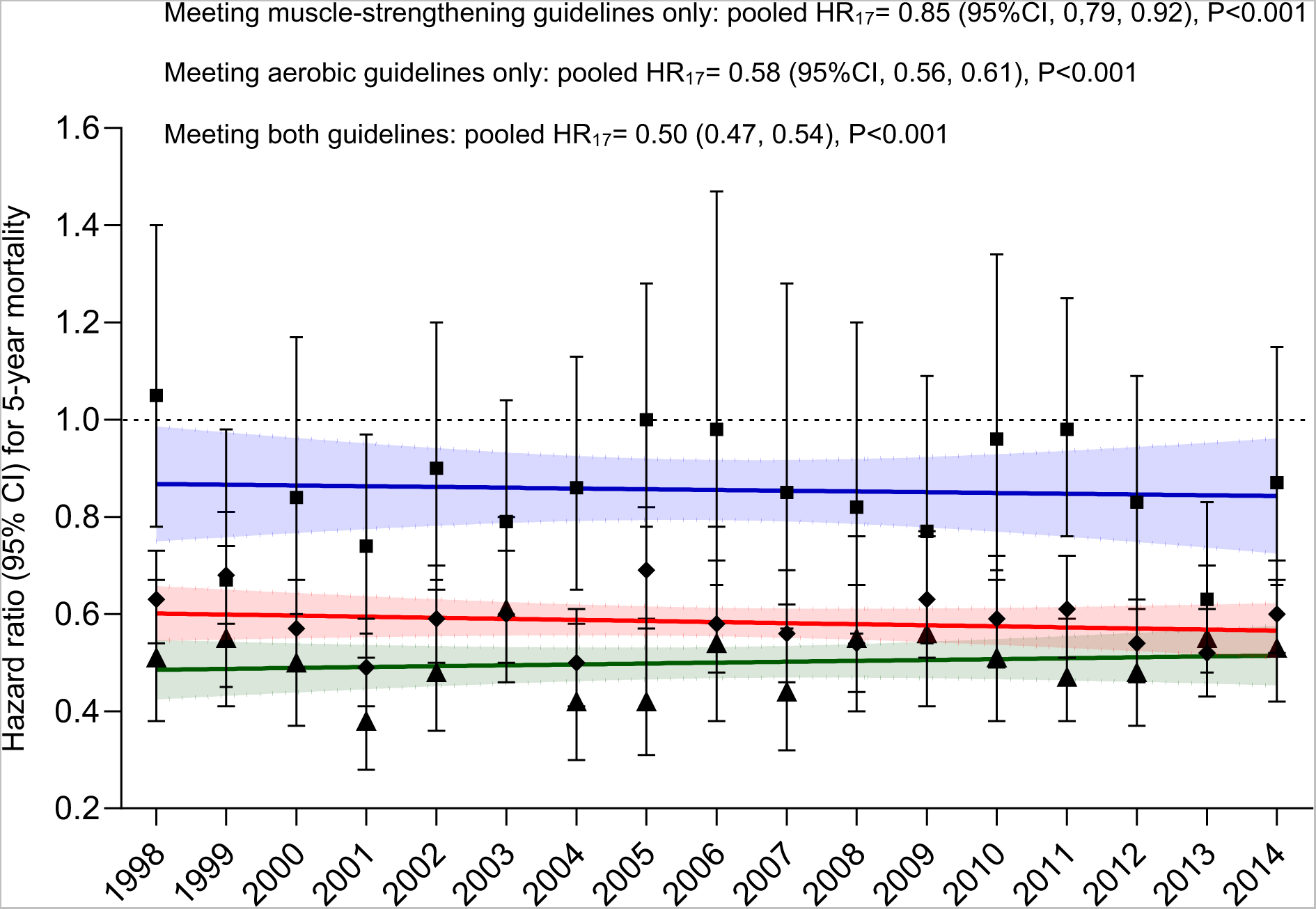
Dose-response associations between combined categories of meeting physical activity guidelines and 5-year mortality in the National Health Interview Surveys 1998-2014. Pooled analysis included 17 annual surveys (1998-2014) as a unique cohort. Trend analysis was summarized with linear regression estimates (standardized B) that used the hazard ratio and cohort year as dependent and independent variables, respectively. The color lines indicate lineal regressions and its 95%CI (all P for trend non-significant). Meeting muscle-strengthening guidelines only: blue. Meeting aerobic physical activity guidelines only: red. Meeting both aerobic and muscle-strengthening guidelines: green. Hazard ratios were adjusted for sex, age, race, education, marital status, chronic conditions, cardiometabolic risk factors, body mass index, smoking and alcohol consumption.

**Figure S2.**
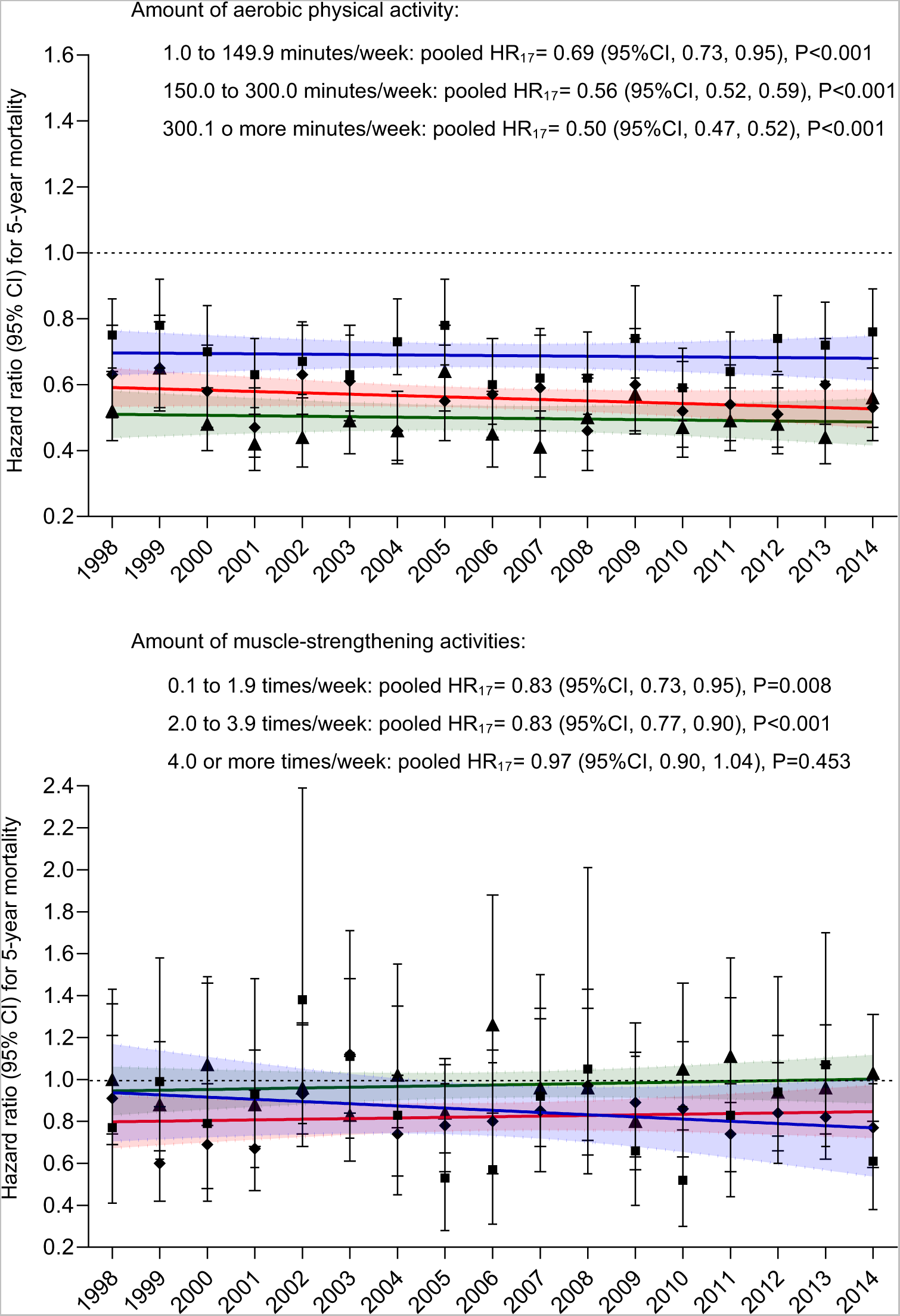
Dose-response associations between amounts of aerobic and muscle-strengthening physical activities and 5-year mortality in the National Health Interview Surveys 1998-2014. Pooled analysis included 17 annual cohorts (1998-2014) as a unique cohort. Trend analysis was summarized with linear regression estimates (standardized B) that used the hazard ratio and cohort year as dependent and independent variables, respectively. The color lines indicate lineal regressions and its 95%CI (all P for trend non-significant). The increasing categories are identified with blue, red, and green colors, respectively. Hazard ratios were adjusted for sex, age, race, education, marital status, chronic conditions, cardiometabolic risk factors, body mass index, smoking, alcohol consumption, and the other type of physical activity.

**Figure S3.**
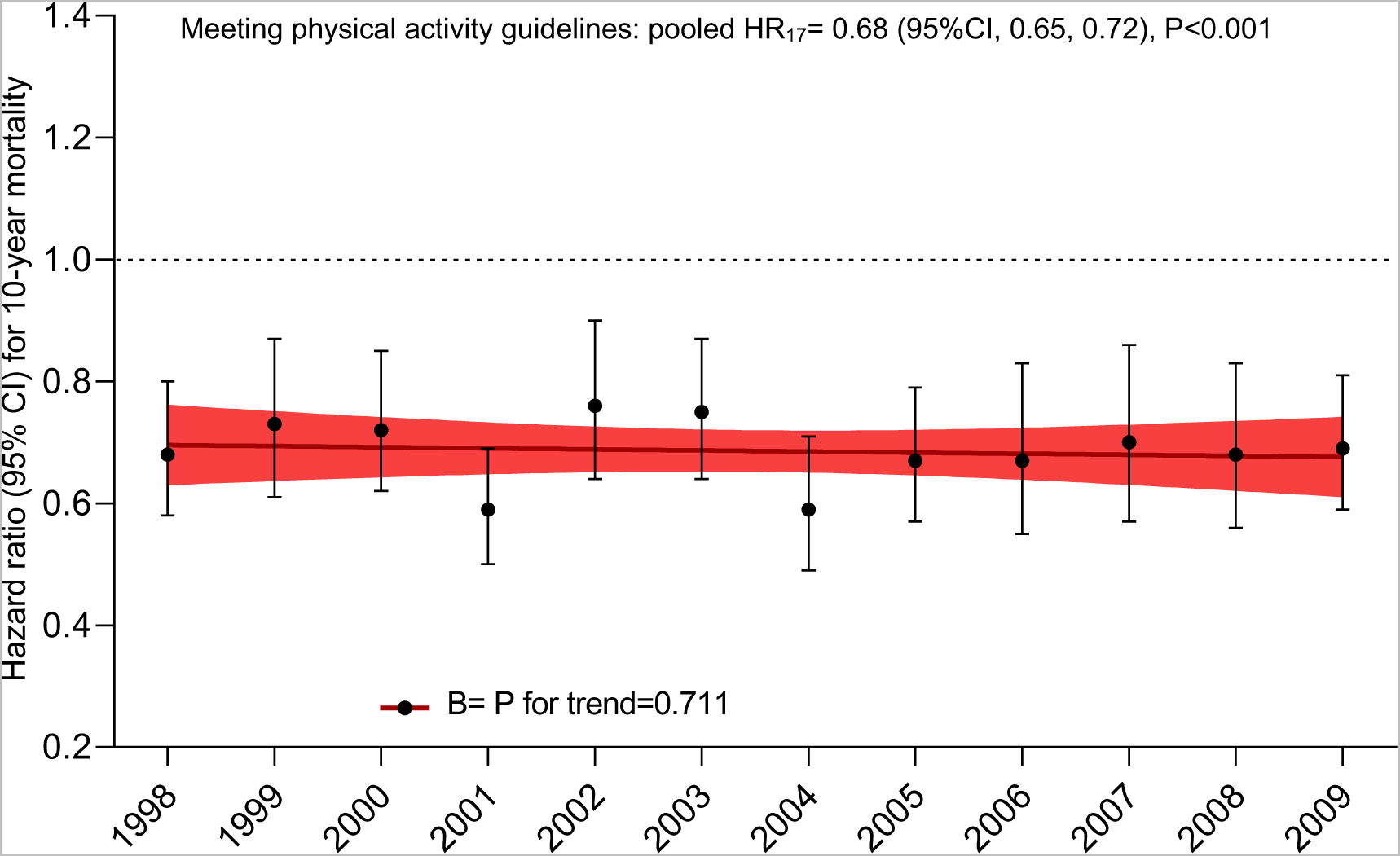
Associations between meeting physical activity guidelines and 10-year mortality in the National Health Interview Surveys 1998-2009. Pooled analysis included 12 annual cohorts (1998-2009). Trend analysis was summarized with linear regression estimates (standardized B) that used the hazard ratio and cohort year as dependent and independent variables. Meeting physical activity guidelines include both recommended aerobic and muscle strengthening activities. The color line indicates lineal regression and its 95%CI. Hazard ratios were adjusted for sex, age, race, education, marital status, chronic conditions, cardiometabolic risk factors, body mass index, smoking, and

